# Investigating the Impact of Caregiver Transportation Needs on Children’s Response to Behavioral and Mental Health Treatment: A Longitudinal Analysis

**DOI:** 10.1101/2021.03.30.21254649

**Authors:** Elizabeth N. Riley, Olga A. Vsevolozhskaya, Dmitri V. Zaykin, Stephen M. Shimshock, John S. Lyons

## Abstract

Despite a rich and developing research literature on the relationships between transportation and healthcare outcomes, the impact of unmet transportation needs on children, particularly on their general psychosocial functioning, is less well understood. We hypothesized that caregiver’s transportation needs may be a key point of intervention for child-level health outcomes, such that the resolution of caregiver transportation needs may have an important, downstream impact on other areas of the child’s functioning. We tested this hypothesis in a sample of children (n=4341) served in a large, statewide public behavioral healthcare system. We conducted a retrospective longitudinal analysis of children served in the system between March 2019 and March 2020, using a specialized assessment tool that captures a comprehensive range of psychosocial functioning: the Child and Adolescent Needs and Strengths (CANS; Lyons, 2009) assessment. Linear mixed-effects analyses demonstrated that, if caregivers’ transportation needs were resolved during the episode of care, those children demonstrated greater improvements on both need reduction and strength building across multiple domains than did their peers whose caregivers had unresolved transportation needs. By the end of the episode of care, children whose caregivers had resolved transportation needs were not significantly different than those children whose caregivers never had transportation needs. The resolution of transportation needs may be an important focus for providers working with children with a complex behavioral health, such that the resolution of these transportation needs seems to have a large, positive downstream impact on their overall functioning.

## 1. Introduction

Over the past several decades, a rich literature has developed seeking to better understanding the relationship between transportation access and healthcare access and outcomes. According to a report from the US Transportation Research Board, an estimated 3.6 million Americans missed or delayed medical care because of a lack of access to transportation in a given year (Transportation Research Board, 2005). Transportation access barriers has been demonstrated to impact utilization of check-ups and ongoing care for chronic conditions, particularly among patients with fewer socioeconomic resources (Arcury, Preisser, Gesler, & Powers, 2005; Syed, Gerber, & Sharp, 2013). This research has studied women, older adults, chronically ill patients, individuals belonging to racial or ethnic minority groups, and adults in rural areas, all of whom represent vulnerable populations for whom transportation access barriers are likely to have a profound impact on healthcare access (c.f., Guidry, Aday, Zhang, & Winn, 1997; Solomon, Wing, Steiner, & Gottlieb, 2020; R. Wallace, Hughes-Cromwick, Mull, & Khasnabis, 2005).

Individuals with transportation difficulties often have multiple and complex medical needs, and lack of or delays in care can accumulate and lead to worse health outcomes (Syed et al., 2013). There is also evidence that reduced transportation access influences more indirect, nonmedical aspects of general health and quality of life, including activities and services related to general well-being, like employment, grocery shopping, and social events (Lee & Sener, 2016; Solomon et al., 2020). The economic impact of transportation barriers and health are notable: federal programs (primarily Medicaid) spent approximately $1.3 billion on nonemergency medical transportation (NEMT) services in 2012 alone, programs that are crucial to allowing many individuals to access their needed direct healthcare services (Solomon et al., 2020).

Despite the dedication of federal and state resources aimed at improving barriers to healthcare access, impaired access due to transportation concerns remains a problem in the United States (US Department of Health and Human Service, 2010). Furthermore, while the use of public transportation is highly beneficial to many with transportation needs, one of the best predictors of improved access to healthcare remains access to a private automobile within the family unit or within one’s immediate social network, particularly as public transportation may not be as readily available in rural areas (Arcury et al., 2005; Mofidi, Rozier, & King, 2002; Yang, Zarr, Kass-Hout, Kourosh, & Kelly, 2006). Although research on the relationships between transportation access and healthcare outcomes continues to develop, the impact of unmet transportation needs of adult caregivers on child level outcomes has received less attention.

### 1.1 Transportation needs and healthcare access in children

While the majority of research on transportation barriers and healthcare access has focused on adult patients, the literature on transportation needs and their impact on children’s healthcare access is nascent. Approximately 9% of children in families with annual incomes less than $50,000 miss essential medical appointments due to transportation barriers, regardless of their insurance status (R. Wallace et al., 2005; Zogby et al., 2001). Despite an urgent and ongoing national conversation about the importance of expanding access to healthcare by increasing the number of children and families with health insurance, it seems clear from the literature that insurance coverage alone is not enough to guarantee healthcare access, nor, more importantly, improved health. Furthermore, many children who are eligible for free transportation through federally- and state-funded transportation programs, still have unmet transportation needs (Yang et el., 2006).

Unmet transportation needs affect children’s access to care across the health spectrum, including issues with obtaining medication, accessing dental care, immunizations, chronic illness care, specialized care, and follow-up emergency care (Smith, Highstein, Jaffe, Fisher, & Strunk, 2002; A. Wallace, Scott, Klinnert, & Anderson, 2004; Yang et al., 2006). One study reported that over half of families cited transportation problems (no ride or no car) as the primary reason for missed appointments for their children (Pesata, Pallija, & Webb, 1999). There may be issues with respect to transportation that are specific to the healthcare and psychosocial needs of children and youth. For example, it appears that children who lack access to medical care due to transportation barriers are more concentrated in urban areas (Wallace et al., 2005). It is also likely that the barriers to healthcare access due to transportation needs are more prevalent in already traditionally underserved populations, such as children belonging to racial or ethnic minority groups (Flores, Abreu, Olivar, & Kastner, 1998; Flores & Tomany-Korman, 2008; Guidry et al., 1997; Mendes, 2016), children with undocumented or vulnerable citizenship status (Weathers, Minkovitz, O’Campo, & Diener-West, 2004), children belonging to the LGBTQ+ community, children with complex medical and psychosocial needs, children whose caregivers lack adequate financial or social resources or support, or children whose caregivers carry a high physical or mental health burden. Much of the research on the relationship between health and transportation needs in children has focused on issues of direct access to care, such as missed appointments or difficulty accessing medication; far less work has been done investigating the impact of caregiver’s transportation needs on children’s overall health and psychosocial functioning over time.

### 1.2 Children served by public systems: Specialized assessment

Hundreds of thousands of children are served by public mental and behavioral health systems each year, and many children served in these systems have highly varied needs and strengths, including a complex presentation of medical, psychological, and psychosocial needs. Furthermore, understanding the nature and impact of caregiver’s needs is a key component of understanding children’s needs and planning for treatment and care within these systems.

Comprehensive assessment for children in this population can be a challenging: in addition to having complex presentations, children served in these systems are frequently under the care of multiple providers. This degree of complexity necessitates the use of a specialized comprehensive assessment strategy in order to fully capture the range of needs and strengths for the child and family. One such tool, the Child and Adolescent Needs and Strengths (CANS; (J. S. Lyons, 2009)), is a non-diagnostic, functional and clinical assessment that utilizes a consensus-building approach focused on a shared vision of the child or youth in care. It is the mostly widely used functional assessment for children and adolescents served in the public sector in the United States, with more than 40 states having statewide implementations in at least one sector. The CANS tool is grounded in qualitative research and is a collaborative, person-centered assessment designed to integrate various stakeholder and provider perspectives, including the perspective of the children, youth and families being served in the system. This assessment leverages a consensus-based process, called post-triangulation measurement (Obeid & Lyons, 2010), which allows for both the inclusion of multiple perspectives (triangulation) into a single metric and the translation of traditionally qualitative data into a scored instrument. The CANS has demonstrated adequate reliability when used by both researchers and clinicians, even at the individual item level (Anderson, Lyons, Giles, Price, & Estle, 2003; Chng, Yip, Pek, Ting, & Chu, 2019; Lyons, Rawal, Yeh, Leon, & Tracy, 2002).

With the CANS, children and youth are rated in areas of life domain functioning, behavioral and emotional needs, risk behaviors, strengths, caregiver resources, and may include other domains/modules that are relevant to a particular jurisdiction (e.g., additional items related to fire setting or suicide; (J. Lyons & Lyons, 2004)). Because it was designed to capture large amounts of data representing the varied and interconnected needs and strengths of children and caregivers served in public systems, the CANS assessment has over 140 potential items rated for each youth. Each item is a single-point assessment of the child or caregiver’s functioning in a particular area (e.g., living situation, community involvement, transportation needs, depression, and cultural identity). Due to the large number of items in the CANS assessment, there will likely be great value in methodological work that seeks to identify “key items” within the larger assessment, the intervention on which can predict and drive change in other areas. Finding areas of intervention with significant downstream impact may be of particular interest for practitioners and administrators in systems that serve client populations with highly varied and complex needs and strengths profiles, like public mental and behavioral health systems.

### 1.3 The current study

Transportation access is a key component of healthcare access in adults and children, and transportation barriers have been shown to negatively impact healthcare outcomes in adults. It is not yet well understood, though, how the transportation needs of adult caregivers impact children’s global health and psychosocial functioning outcomes. Many children in this country are served in complex behavioral healthcare systems. In these systems, it is often challenging to know which area of intervention will have the greatest downstream impact on other areas of functioning. Due to the importance of transportation access to health and healthcare in adults, we hypothesize that caregiver transportation needs may be a key point of intervention for child-level health outcomes. We further hypothesize that the resolution of caregiver transportation needs may have large downstream impact on other areas of the child’s functioning. We tested this hypothesis in a sample of children served in a large, public behavioral and mental healthcare system in order to determine whether the presence, and ultimately the resolution, of caregivers’ transportation needs predicted change in other areas of global health and psychosocial functioning for their child over the course of treatment.

## 1. Methods

### 2.1 Design and sample

We conducted a retrospective longitudinal analysis of data from the ICANS database, which stores and manages the Child and Adolescent Needs and Strengths (CANS) assessments from the Idaho Department of Behavioral Health (Idaho Deprtment of Health & Welfare, 2020). The study sample initially included all participants aged ≤ 20 years that entered the publicly funded Mental and Behavioral Health System in Idaho between March 2019 and March 2020 (*N* = 7919). Participants with assessments after March 2020 were excluded from the analysis because of concerns of potential sample differences, particularly with respect to transportation needs due to the COVID-19 pandemic. Because this study was a retrospective longitudinal analysis, we further excluded those with a single CANS assessment (*N* = 3380), multiple re-entrees into the system (i.e., youth and children that exited the mental healthcare system and then re-entered it within the study period; *N* = 66), children less than 5 years old (*N* = 99), and participants with missing gender information (*N* = 33). The total remaining sample size included 4341 participants (2042 girls and 2326 boys) with a mean age of 12.2 (SD = 3.67) and with an average 3.05 assessments for girls and 3.11 assessments for boys. This research was approved by the University of Kentucky Institutional Review Board.

### 2.2 Measures

While in the Idaho Mental and Behavioral Health System, participants are assessed using the CANS on areas where they demonstrate strength and areas where they need support. The Idaho CANS uses 82 items across six domains: strengths (16 items), life-functioning (14 items), behavioral/emotional needs (18 items), risk behaviors (15 items), and caregiver resources (19 items). The full list of CANS items adopted by Idaho’s Department of Behavioral Health is available in an online reference guide (Idaho Department of Health & Welfare, 2017).

For ratings of needs, the CANS instrument scores each item on a 0 to 3 scale: 0 = no need for remedial action; 1 = watchful waiting to see whether remedial action is necessary; 2 = remedial action is necessary; 3 = remedial action immediately or intensive remedial action needed. For ratings of strengths, the scoring is: 0 = centerpiece strength used as the focus or foundation of a strength-based plan; 1 = useful strength used in a strength-based plan; 2 = strengths have been identified but they require significant building efforts before they can be effectively utilized; 3 = efforts needed in order to identify this as a potential strength (J. S. Lyons, 2009). For both needs and strengths, items that score 2 or 3 are called “actionable items,” indicating an area for targeted clinical intervention, and items that score 0 or 1 are considered “non-actionable.” In this study, items were scored as binary based on whether or not they were considered “actionable” (non-actionable = 0, actionable = 1).

#### 2.2.1 Dependent variables

Our two primary outcomes of interest were two proportions: (1) the proportion of needs resolved over needs acquired during the study timeframe, calculated for the life-functioning, behavioral/emotional needs, and risk behaviors domains, and (2) the proportion of strengths built over strengths to develop during the study timeframe among children and youth in Idaho Mental and Behavioral Health System. Items from the caregiver resources domain were used to adjust for potential confounders in our model, as described below.

To calculate the proportion of needs resolved over needs acquired during the study timeframe, we first calculated the total number of actionable need-related items “acquired up to date.” For example, suppose we are calculating the total number of actionable need-related items “acquired up to date” in the risk behavior domain (total number of items = 15). If at the baseline assessment, a study participant had items 6, 9, and 13 scored as actionable and the rest of the items scored as non-actionable, the total number of actionable items acquired up to date will be equal to 3. If at the second assessment, the same participant had items 1, 6 and 13 scored as actionable, with the rest of the items scored as non-actionable. Then, the total number of actionable items acquired up to date will be equal to 4, because the study participant has accumulated one additional new actionable item (item 1) in addition to the three items that were actionable during the first assessment (items 6, 9, and 13). Repeat actionable items across assessments are only counted once. Figure 1 illustrates the total number of actionable items acquired up to date among our study participants across the life-functioning, behavioral/emotional needs, and risk behaviors CANS domains.

**Figure 1.**
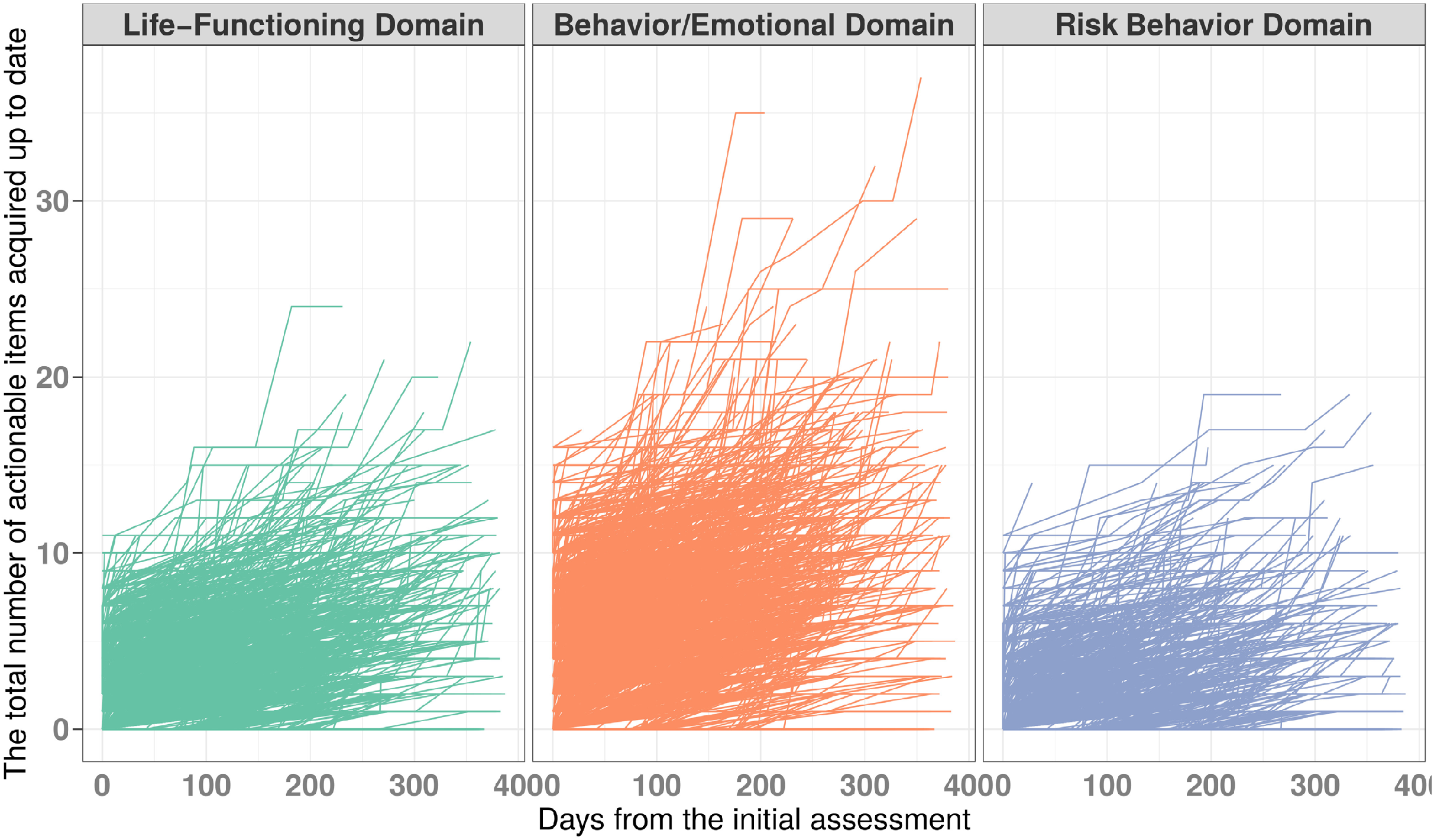
Number of actionable needs acquired during the study timeframe for all participants across all three CANS needs domains. *Note*. Each line represents the number of needs acquired (as a cumulative and increasing score) for each individual study participant throughout their care episode.

Then, we calculated the total number of need-related actionable items that were “resolved up to date.” At initial assessment, all subjects would have zero items resolved. If previously actionable items become non-actionable in subsequent assessments, those were considered “resolved items.” In the example above, the participant has 0 items resolved on the first assessment and one item resolved (item 9) on the second assessment. Figure 2 illustrates the total number of actionable items “resolved up to date” among our study participants across CANS domains. Finally, to calculate the proportion of needs resolved over needs acquired during the study timeframe we scaled the total number of actionable items resolved up to date by the total number of actionable items accumulated by the last assessment. Figure 3 illustrates our constructed feature of the proportion of needs resolved over needs acquired during the study timeframe among our study participants across the CANS domains.

**Figure 2.**
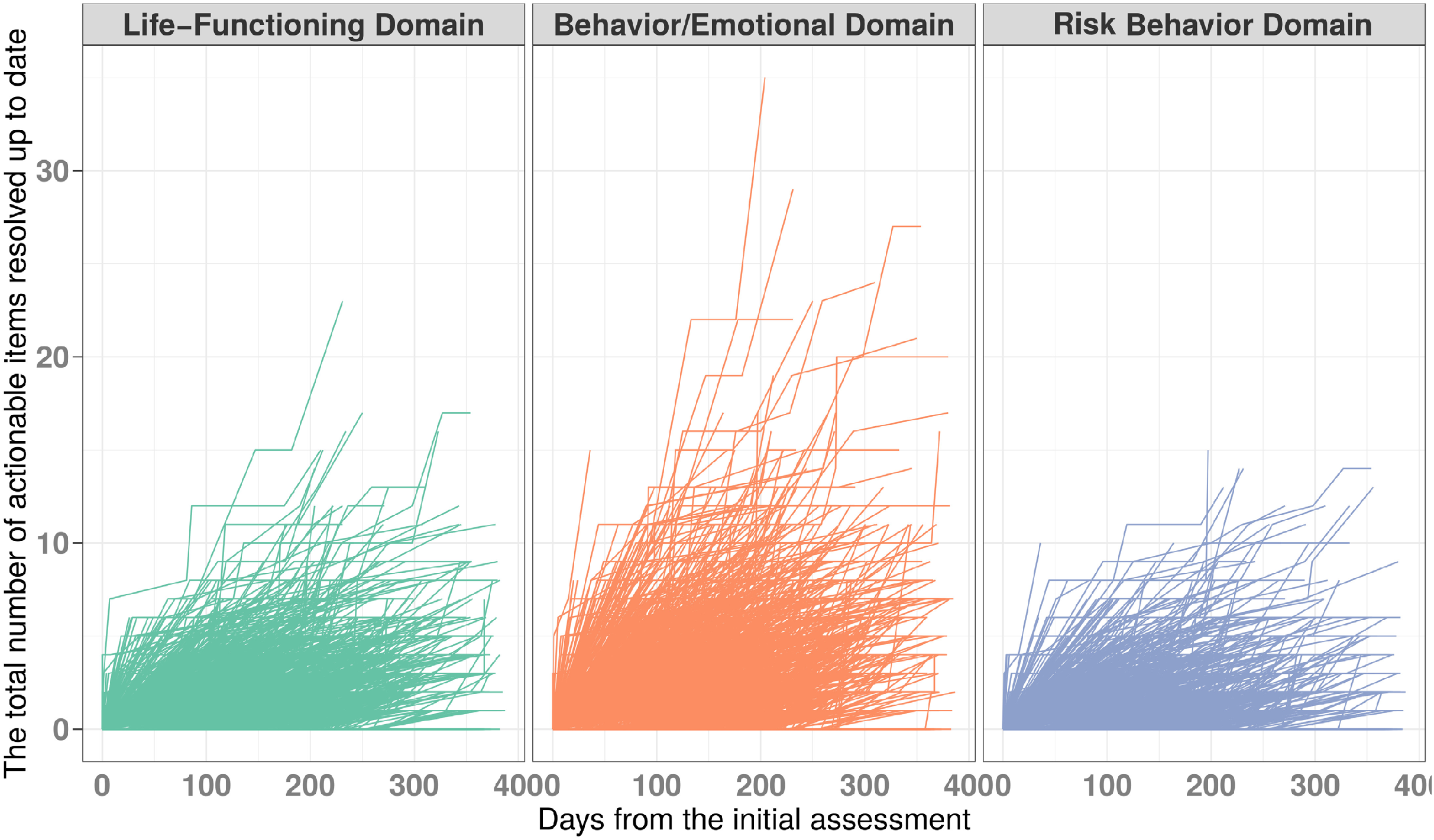
Number of actionable needs resolved during the study timeframe for all participants across all three CANS needs domains. *Note*. Each line represents the number of needs resolved (as a cumulative and increasing score) for each individual study participant throughout their care episode.

**Figure 3.**
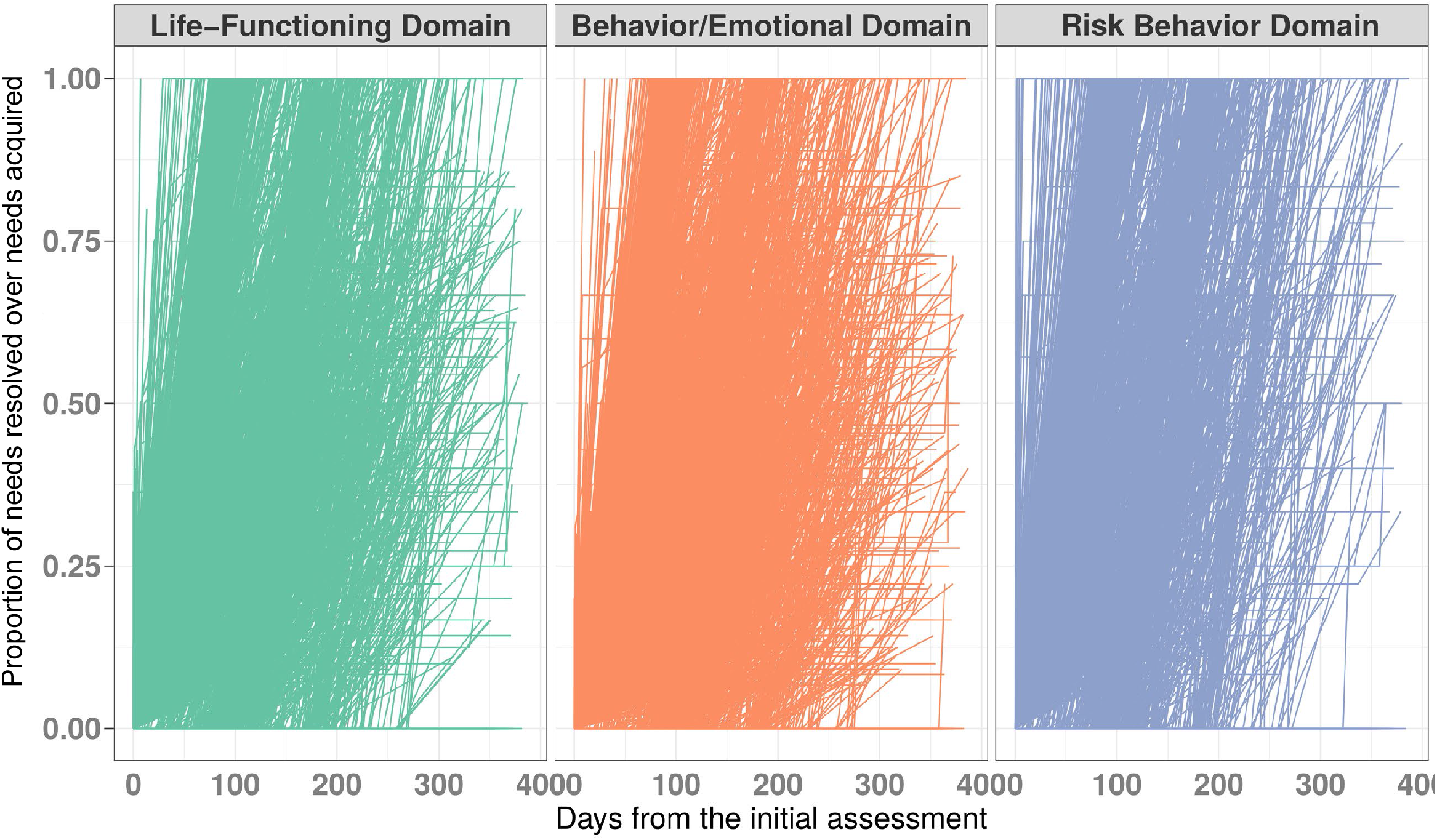
Proportion of needs resolved over needs acquired during the study timeframe for all participants across all three CANS needs domains. *Note*. Each line represents the proportion of needs resolved (relative to needs acquired) for each individual study participant throughout their care episode. If a participant resolved all acquired needs, the proportion of needs resolved over needs acquired would be 1.0.

The proportion of strengths built over strengths to develop during the study timeframe was calculated using a similar process, constructed as strengths acquired over strengths to develop for each participant during the study timeframe.

#### 2.2.2 Explanatory and confounding variables

The primary independent variable of interest was the change in the transportation needs of the caregiver constellation for a child at the time of care. This caregiver constellation includes all individuals who are considered caregivers for the child and is not limited to rating a single caregiver. This item rates the level of transportation required to ensure that a child/youth can effectively participate in his/her treatment. If the transportation item was never rated as actionable, across all CANS assessments, then we defined this participant as having “no issues” with transportation needs during the study timeframe. If this item was actionable during one assessment but became non-actionable by the last assessment, we defined this as a “resolved” transportation need. If the transportation item remained actionable by the last assessment, then we defined the transportation need as being “unresolved” for this study participant. Participants were grouped based on these three levels of transportation needs: no issues, resolved, or unresolved.

To adjust for confounders associated with socioeconomic status, financial resources, and general patterns of caregiver’s behavior, we calculated first five principal components from items in the caregiver resources domain (that explained 53% of variation) and used them as another set of explanatory variables to adjustment for potential confounding associated with other caregiver’s characteristics beyond unmet transportation needs. We also included individual characteristics of children and youth, such as age, gender, and race/ethnicity in our model.

### 2.3 Statistical analysis

We calculated descriptive statistics for participants at different levels of caregiver’s transportation need (no issues, resolved, or unresolved). Then, we used a linear mixed-effects model to yield estimates of changes in participant’s domain-specific proportion of needs resolved over needs acquired during the study timeframe and the proportion of strengths built over strengths to develop during the study timeframe by the level of caregiver’s transportation need.

All models also included participant’s length of stay in the system (i.e., number of days from the initial assessment), gender, race/ethnicity, and the first five principal components of caregiver’s characteristics as the fixed effects. The random effects included a random intercept and a random slope for the length of stay in the system. All analyses were conducted using R v. 3.6.3 statistical software and statistical significance was set at *P* < 0.05.

## 2. Results

Of the 4341 participants, 4033 reported no transportation needs during the study timeframe, 124 reported transportation needs as resolved, and 184 as unresolved. Descriptive data in Table 1 shows the patterns of children’s psychosocial functioning outcomes relative to caregiver’s transportation needs during the study timeframe. These data suggest that youth with caregivers who experienced transportation access needs at some point during the study timeframe had a higher number of overall needs to resolve, across multiple domains, and a higher number of overall strengths to develop compared to youth whose caregivers had no transportation needs. Furthermore, children and teenagers with resolved transportation needs demonstrated a higher number of strengths acquired and needs resolved up to date, compared to those participants with unresolved transportation needs during the study timeframe.

**Table 1.**
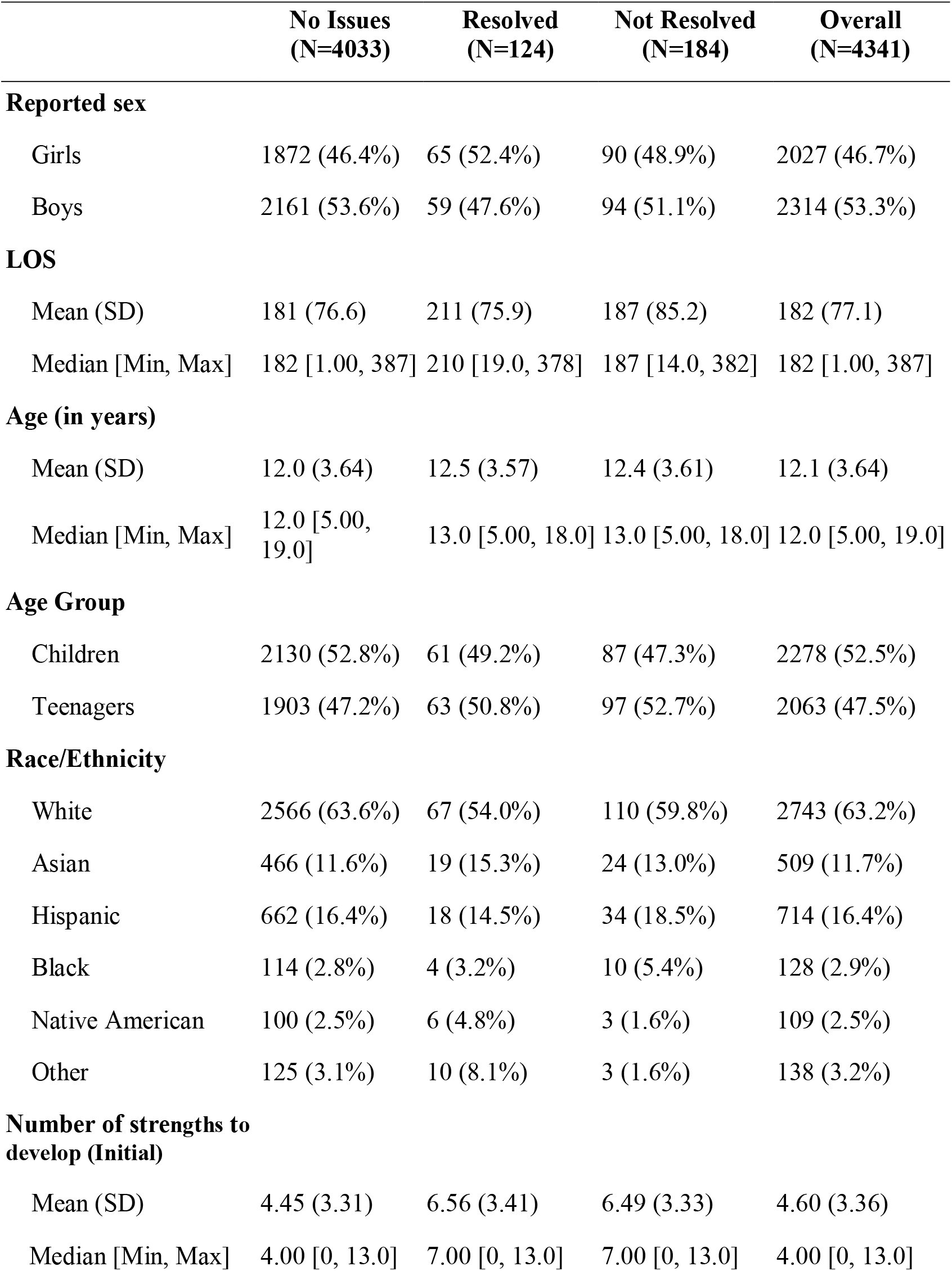
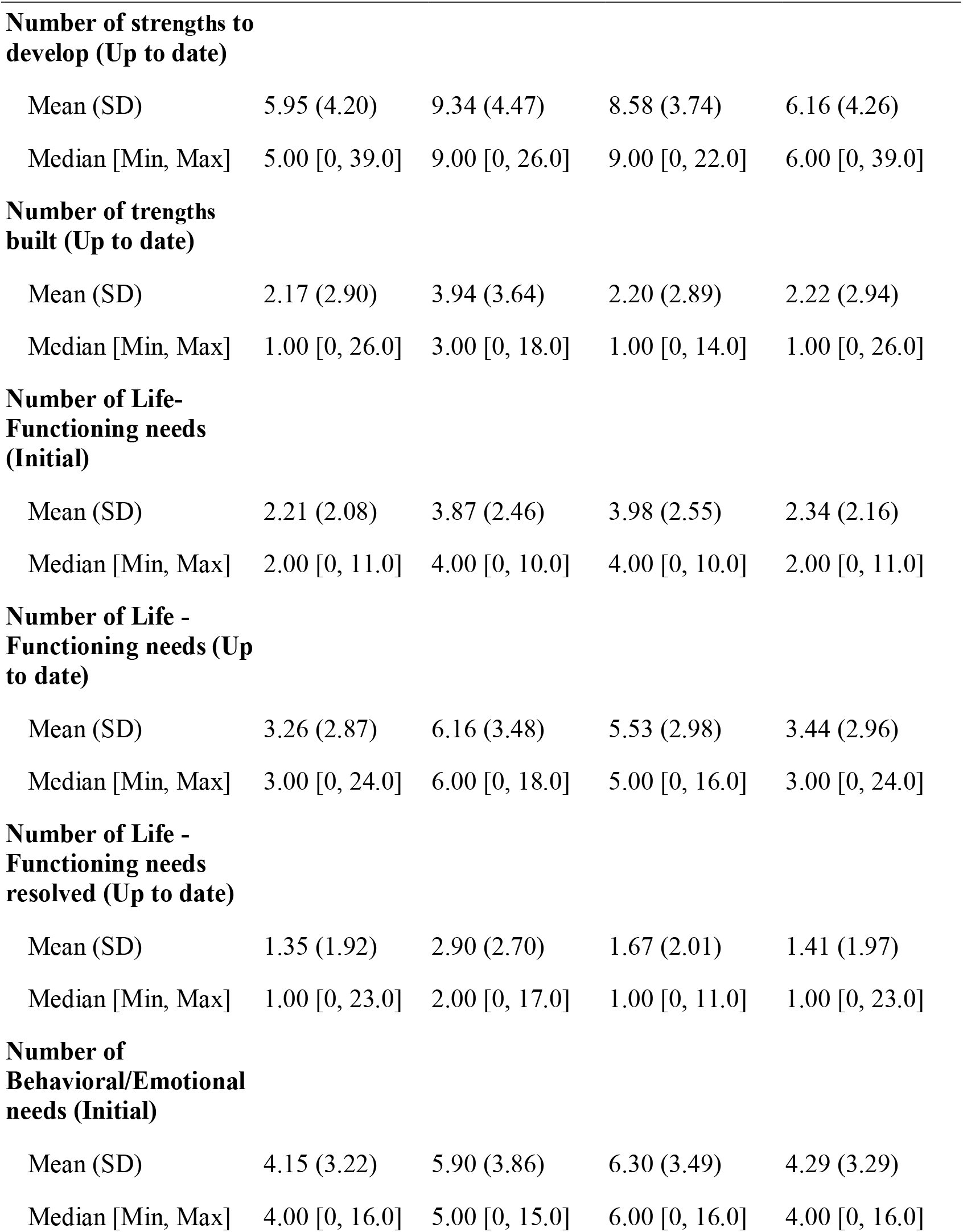
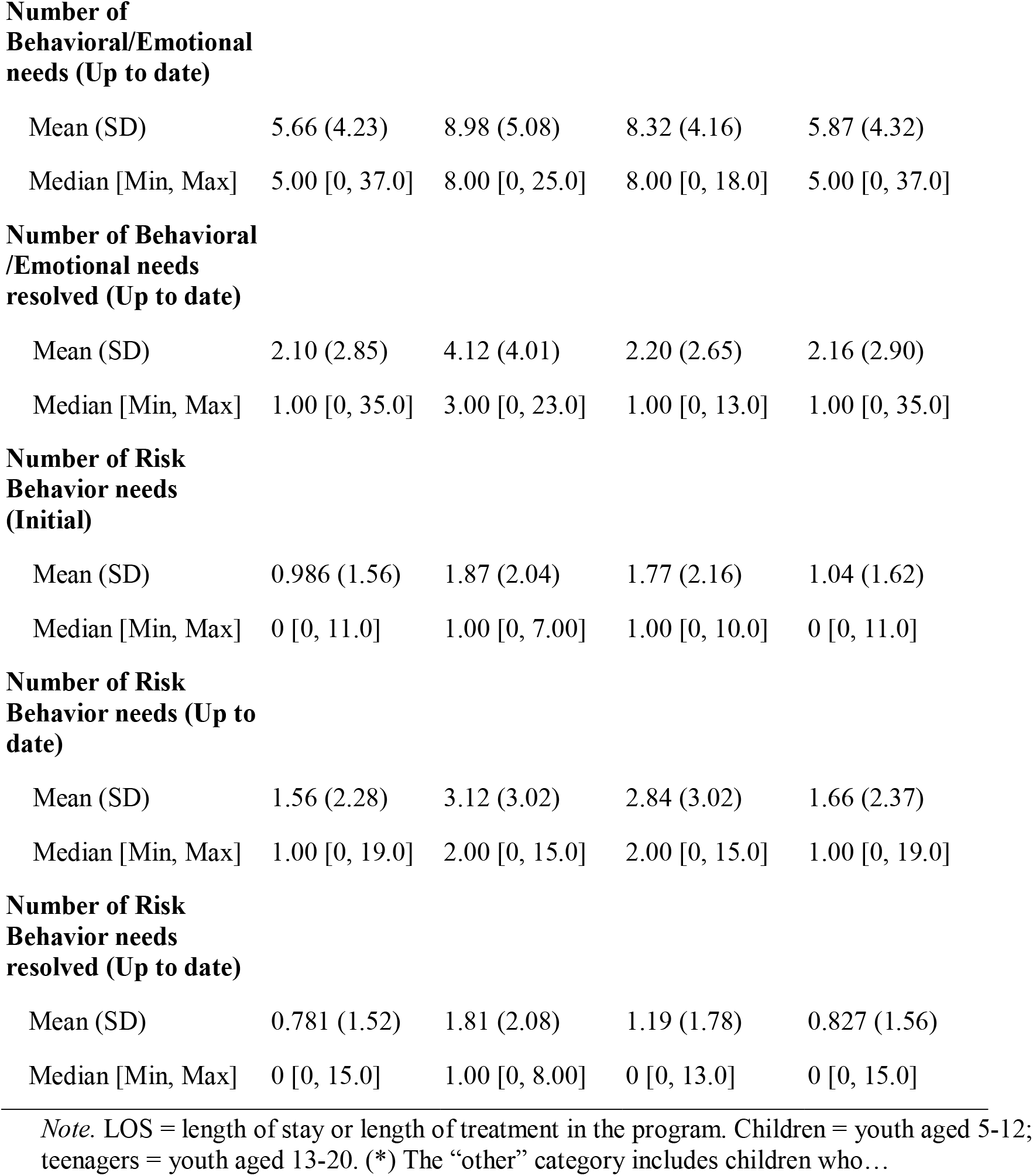
Characteristics of study participants from the Idaho Behavioral Health System, ages 5-19, based on level of caregiver transportation needs (no issues, resolved, not resolved).

|  | **No Issues**<br>**(N=4033)** | **Resolved**<br>**(N=124)** | **Not Resolved**<br>**(N=184)** | **Overall**<br>**(N=4341)** |
| --- | --- | --- | --- | --- |
| **Reported sex** |  |  |  |  |
| Girls | 1872 (46.4%) | 65 (52.4%) | 90 (48.9%) | 2027 (46.7%) |
| Boys | 2161 (53.6%) | 59 (47.6%) | 94 (51.1%) | 2314 (53.3%) |
| **LOS** |  |  |  |  |
| Mean (SD) | 181 (76.6) | 211 (75.9) | 187 (85.2) | 182 (77.1) |
| Median [Min, Max] | 182 [1.00, 387] | 210 [19.0, 378] | 187 [14.0, 382] | 182 [1.00, 387] |
| **Age (in years)** |  |  |  |  |
| Mean (SD) | 12.0 (3.64) | 12.5 (3.57) | 12.4 (3.61) | 12.1 (3.64) |
| Median [Min, Max] | 12.0 [5.00, 19.0] | 13.0 [5.00, 18.0] | 13.0 [5.00, 18.0] | 12.0 [5.00, 19.0] |
| **Age Group** |  |  |  |  |
| Children | 2130 (52.8%) | 61 (49.2%) | 87 (47.3%) | 2278 (52.5%) |
| Teenagers | 1903 (47.2%) | 63 (50.8%) | 97 (52.7%) | 2063 (47.5%) |
| **Race/Ethnicity** |  |  |  |  |
| White | 2566 (63.6%) | 67 (54.0%) | 110 (59.8%) | 2743 (63.2%) |
| Asian | 466 (11.6%) | 19 (15.3%) | 24 (13.0%) | 509 (11.7%) |
| Hispanic | 662 (16.4%) | 18 (14.5%) | 34 (18.5%) | 714 (16.4%) |
| Black | 114 (2.8%) | 4 (3.2%) | 10 (5.4%) | 128 (2.9%) |
| Native American | 100 (2.5%) | 6 (4.8%) | 3 (1.6%) | 109 (2.5%) |
| Other | 125 (3.1%) | 10 (8.1%) | 3 (1.6%) | 138 (3.2%) |
| **Number of strengths to develop (Initial)** |  |  |  |  |
| Mean (SD) | 4.45 (3.31) | 6.56 (3.41) | 6.49 (3.33) | 4.60 (3.36) |
| Median [Min, Max] | 4.00 [0, 13.0] | 7.00 [0, 13.0] | 7.00 [0, 13.0] | 4.00 [0, 13.0] |

|  | No Issues<br>(N=4033) | Resolved<br>(N=124) | Not Resolved<br>(N=184) | Overall<br>(N=4341) |
| --- | --- | --- | --- | --- |
| **Number of strengths to develop (Up to date)** |  |  |  |  |
| Mean (SD) | 5.95 (4.20) | 9.34 (4.47) | 8.58 (3.74) | 6.16 (4.26) |
| Median [Min, Max] | 5.00 [0, 39.0] | 9.00 [0, 26.0] | 9.00 [0, 22.0] | 6.00 [0, 39.0] |
| **Number of strengths built (Up to date)** |  |  |  |  |
| Mean (SD) | 2.17 (2.90) | 3.94 (3.64) | 2.20 (2.89) | 2.22 (2.94) |
| Median [Min, Max] | 1.00 [0, 26.0] | 3.00 [0, 18.0] | 1.00 [0, 14.0] | 1.00 [0, 26.0] |
| **Number of Life-Functioning needs (Initial)** |  |  |  |  |
| Mean (SD) | 2.21 (2.08) | 3.87 (2.46) | 3.98 (2.55) | 2.34 (2.16) |
| Median [Min, Max] | 2.00 [0, 11.0] | 4.00 [0, 10.0] | 4.00 [0, 10.0] | 2.00 [0, 11.0] |
| **Number of Life - Functioning needs (Up to date)** |  |  |  |  |
| Mean (SD) | 3.26 (2.87) | 6.16 (3.48) | 5.53 (2.98) | 3.44 (2.96) |
| Median [Min, Max] | 3.00 [0, 24.0] | 6.00 [0, 18.0] | 5.00 [0, 16.0] | 3.00 [0, 24.0] |
| **Number of Life - Functioning needs resolved (Up to date)** |  |  |  |  |
| Mean (SD) | 1.35 (1.92) | 2.90 (2.70) | 1.67 (2.01) | 1.41 (1.97) |
| Median [Min, Max] | 1.00 [0, 23.0] | 2.00 [0, 17.0] | 1.00 [0, 11.0] | 1.00 [0, 23.0] |
| **Number of Behavioral/Emotional needs (Initial)** |  |  |  |  |
| Mean (SD) | 4.15 (3.22) | 5.90 (3.86) | 6.30 (3.49) | 4.29 (3.29) |
| Median [Min, Max] | 4.00 [0, 16.0] | 5.00 [0, 15.0] | 6.00 [0, 16.0] | 4.00 [0, 16.0] |
| **Number of Behavioral/Emotional needs (Up to date)** |  |  |  |  |
| Mean (SD) | 5.66 (4.23) | 8.98 (5.08) | 8.32 (4.16) | 5.87 (4.32) |
| Median [Min, Max] | 5.00 [0, 37.0] | 8.00 [0, 25.0] | 8.00 [0, 18.0] | 5.00 [0, 37.0] |
| **Number of Behavioral/Emotional needs resolved (Up to date)** |  |  |  |  |
| Mean (SD) | 2.10 (2.85) | 4.12 (4.01) | 2.20 (2.65) | 2.16 (2.90) |
| Median [Min, Max] | 1.00 [0, 35.0] | 3.00 [0, 23.0] | 1.00 [0, 13.0] | 1.00 [0, 35.0] |
| **Number of Risk Behavior needs (Initial)** |  |  |  |  |
| Mean (SD) | 0.986 (1.56) | 1.87 (2.04) | 1.77 (2.16) | 1.04 (1.62) |
| Median [Min, Max] | 0 [0, 11.0] | 1.00 [0, 7.00] | 1.00 [0, 10.0] | 0 [0, 11.0] |
| **Number of Risk Behavior needs (Up to date)** |  |  |  |  |
| Mean (SD) | 1.56 (2.28) | 3.12 (3.02) | 2.84 (3.02) | 1.66 (2.37) |
| Median [Min, Max] | 1.00 [0, 19.0] | 2.00 [0, 15.0] | 2.00 [0, 15.0] | 1.00 [0, 19.0] |
| **Number of Risk Behavior needs resolved (Up to date)** |  |  |  |  |
| Mean (SD) | 0.781 (1.52) | 1.81 (2.08) | 1.19 (1.78) | 0.827 (1.56) |
| Median [Min, Max] | 0 [0, 15.0] | 1.00 [0, 8.00] | 0 [0, 13.0] | 0 [0, 15.0] |

*Note.* LOS = length of stay or length of treatment in the program. Children = youth aged 5-12; teenagers = youth aged 13-20. (\*) The “other” category includes children who...

Linear mixed-effects analyses revealed that the proportion of needs resolved over needs acquired during the study timeframe among youth whose caregivers resolved their transportation needs was no different from youth whose caregivers never demonstrated transportation needs during the study timeframe. Youth whose caregivers did not resolve transportation issues had a significantly lower proportion of needs resolved over needs acquired during the study timeframe (Table 2); that is, they resolved fewer needs (relative to needs acquired) than those youth whose caregivers did resolve transportation needs. For example: in the behavioral/emotional needs domain, by the midpoint of the study timeframe (i.e., Time centered = 0; approximately day 100), youth with no transportation needs tend to resolve about 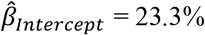 of needs in the behavioral/emotional domain. Compared to the group with no transportation needs, youth with resolved transportation needs tend to resolve 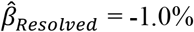 fewer needs (*P* = 0.693), and youth with unresolved transportation needs tend to address 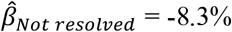 fewer needs (*P* < 0.001). After every additional 90 days in the system (i.e., Time centered = 1), youth with no demonstrated transportation needs tend to resolve an additional 21.6% of behavioral/emotional needs. Relative to the group with no transportation needs, youth with resolved transportation issues tend to resolve −0.9% fewer needs (*P* = 0.692), whereas those with unresolved transportation needs tend to resolve −7.8% fewer needs (*P* < 0.001). Similar pattern are demonstrated by the coefficients for the analyses of the risk behaviors and life-functioning domains (Table 2).

**Table 2.**
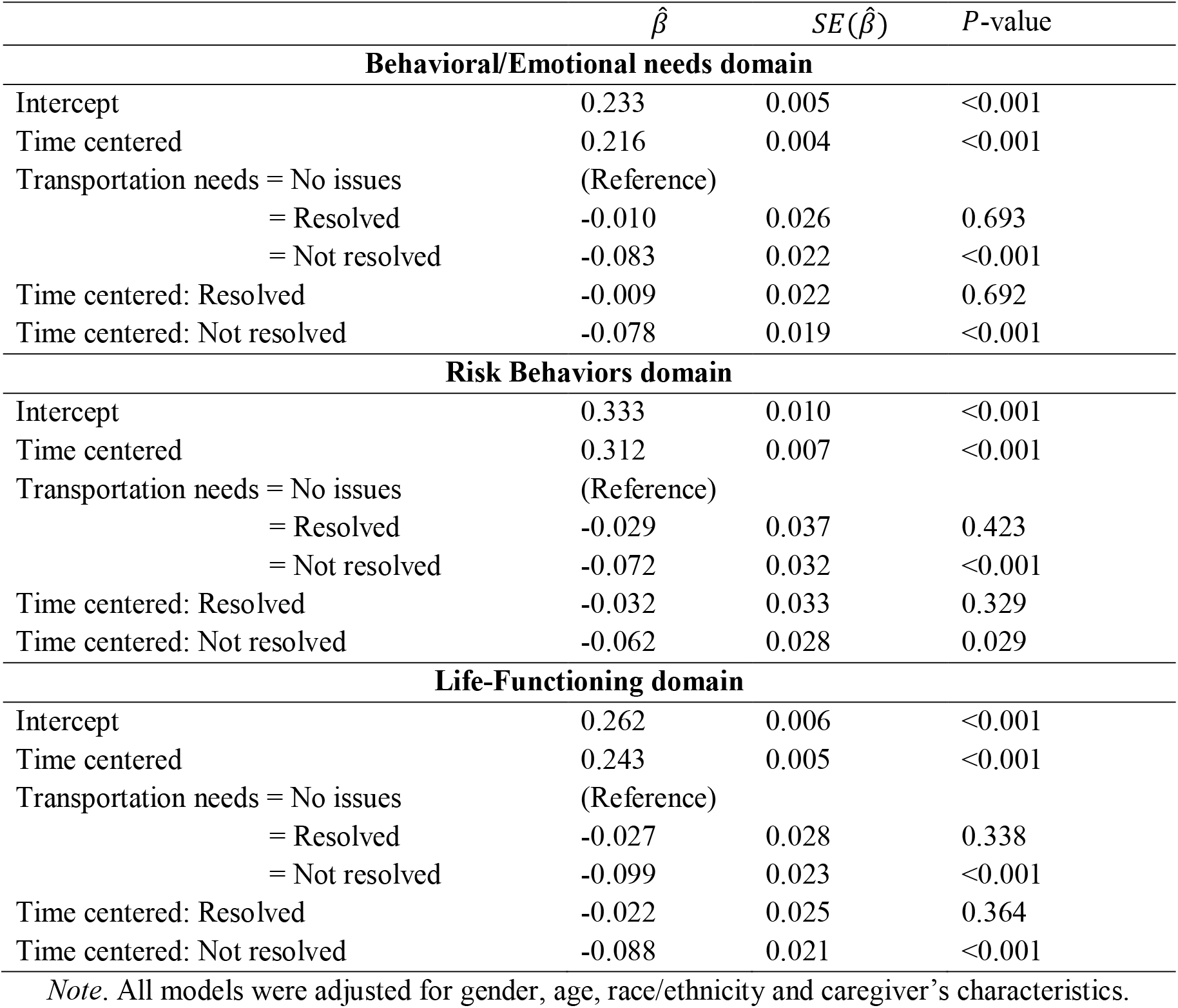
Linear mixed-effects model estimates for the prediction of the proportion of needs resolved over needs acquired during the study timeframe, based on caregiver’s transportation needs.

| | $\hat{\beta}$ | $SE(\hat{\beta})$ | P-value |
| --- | --- | --- | --- |
| **Behavioral/Emotional needs domain** |  |  |  |
| Intercept | 0.233 | 0.005 | <0.001 |
| Time centered | 0.216 | 0.004 | <0.001 |
| Transportation needs = No issues | (Reference) |  |  |
| = Resolved | -0.010 | 0.026 | 0.693 |
| = Not resolved | -0.083 | 0.022 | <0.001 |
| Time centered: Resolved | -0.009 | 0.022 | 0.692 |
| Time centered: Not resolved | -0.078 | 0.019 | <0.001 |
| **Risk Behaviors domain** |  |  |  |
| Intercept | 0.333 | 0.010 | <0.001 |
| Time centered | 0.312 | 0.007 | <0.001 |
| Transportation needs = No issues | (Reference) |  |  |
| = Resolved | -0.029 | 0.037 | 0.423 |
| = Not resolved | -0.072 | 0.032 | <0.001 |
| Time centered: Resolved | -0.032 | 0.033 | 0.329 |
| Time centered: Not resolved | -0.062 | 0.028 | 0.029 |
| **Life-Functioning domain** |  |  |  |
| Intercept | 0.262 | 0.006 | <0.001 |
| Time centered | 0.243 | 0.005 | <0.001 |
| Transportation needs = No issues | (Reference) |  |  |
| = Resolved | -0.027 | 0.028 | 0.338 |
| = Not resolved | -0.099 | 0.023 | <0.001 |
| Time centered: Resolved | -0.022 | 0.025 | 0.364 |
| Time centered: Not resolved | -0.088 | 0.021 | <0.001 |

*Note.* All models were adjusted for gender, age, race/ethnicity and caregiver's characteristics.

Table 3 shows estimated model coefficients for the proportion of strengths built over strengths to develop during the study timeframe by the caregiver’s transportation needs. Based on the data presented in this table, we can conclude that the strength-acquiring proportion among youth whose caregivers resolved their transportation access issues was no different from youth whose caregivers never had transportation needs. However, youth whose caregivers had unresolved transportation needs had a significantly lower proportion of strengths built over strengths to develop during the study timeframe than both the “never transportation needs” or the “resolved transportation needs” groups.

**Table 3.** Linear mixed-effects model estimates for the prediction of the proportion of strengths built over strengths to develop during the study timeframe, based on caregiver’s transportation needs.

| | $\hat{\beta}$ | $SE(\hat{\beta})$ | *P*-value |
| --- | --- | --- | --- |
| **Strengths domain** |  |  |  |
| Intercept | 0.222 | 0.005 | <0.001 |
| Time centered | 0.200 | 0.004 | <0.001 |
| Transportation needs = No issues | (Reference) |  |  |
| = Resolved | -0.013 | 0.023 | 0.590 |
| = Not resolved | -0.089 | 0.019 | <0.001 |
| Time centered: Resolved | -0.008 | 0.021 | 0.661 |
| Time centered: Not resolved | -0.078 | 0.017 | <0.001 |

*Note.* This model was adjusted for gender, age, race/ethnicity and caregiver's characteristics.

## 3. Discussion

The results of this study indicate that caregiver transportation needs are associated with higher levels of children’s psychosocial “needs to resolve” and “strengths to build” at the outset of treatment. Among all children whose caregivers had identified transportation needs, children belonging to racial and ethnic minority groups, older children, and male-identified children present to treatment with more needs and fewer strengths than their white, younger, and female-identified peers. However, if caregiver transportation needs are resolved, there is a significant, positive downstream impact: children whose caregiver’s transportation needs were resolved demonstrated greater strength building proportions (relative to strengths to develop) and greater need reduction proportions (relative to needs acquired) than did their peers whose caregivers had unresolved transportation needs. By the end of the study period, those children with caregivers whose transportation needs were resolved were not significantly different than those children whose caregivers never had transportation needs. While the presence of transportation barriers is clearly impactful, it seems that the resolution of these needs may be even more impactful: the resolution of transportation needs may be a crucial area of clinical focus for providers treating children with a variety of psychosocial needs and strengths served by complex systems. If transportation needs are not resolved, this could be a limiting factor for treatment; without the resolution of transportation needs during treatment, one might expect less improvement in other areas of functioning for this child over time.

There is a small but growing literature on how transportation barriers impact the health and healthcare outcomes of children, but much of this research focuses on specific metrics of healthcare access, such as appointment attendance. To our knowledge, this study was the first to examine the impact of caregiver transportation needs on a broad range of children’s psychosocial needs and strengths while in treatment. It was furthermore the first investigation into how the resolution, or irresolution, of caregiver transportation needs impacted the development of children’s psychosocial needs and strengths over time, while engaged in mental or behavioral health treatment. Finally, this study represents an important methodological advance in considering and analyzing data from complex systems: we used a system-specific assessment tool designed to capture a wide range of psychosocial functioning based on communimetric and consensus-building techniques. This methodological approach was an attempt to identify key items within the larger assessment, the intervention on which has a significant and far-reaching downstream impact.

The stability and magnitude of the present findings are striking. The data used in this study are unique and highly complex: the data are comprised of several thousand cases, each case with varying numbers of assessments over the 6-month study period, each assessment with varying numbers of data points within each assessment. The fact that caregiver transportation need resolution was consistently identified as a significant predictor of change in child-level outcomes, across multiple domains of functioning, speaks to the robustness of this effect for the present sample. Due to the large number of items in the CANS assessment, identifying “key items” within this larger assessment where intervention can predict and drive change in other areas may aid in clinical and administrative decision support. Identifying key items of the CANS, the intervention on which can significantly affect functioning in other areas, would be a methodological and analytic development of great public health value to systems that serve children with complex presentations of medical needs, psychological needs, psychosocial needs/strengths, and caregiver needs (Cordell, Snowden, & Hosier, 2016). Based on the results of this study, we suggest that caregiver transportation needs may be one of those key impactful items, particularly for clients with complex needs and strengths profiles. Complex healthcare systems, such as public behavioral health systems, are often overtaxed and underfunded; the ability to identify key predictive items that drive change in other areas may aid in clinical and administrative decision support.

### 4.1 Limitations and future directions

This study is, of course, not without limitations. First, though the overall sample was large, the study sample of children whose caregivers had identified transportation needs was relatively small and homogeneous. Given the wealth of literature demonstrating the prevalence and impact of transportation needs among adult caregivers and children identifying as part of racial or ethnic minority group (Flores & Tomany-Korman, 2008; Guidry et al., 1997), analyses prioritizing the study of these structural barriers are a crucial area of research. While we were able to examine some of the differences in presenting needs/strengths patterns by race and ethnicity, we were not able to explore interaction effects of transportation needs by these variables due to small sample sizes of children identifying as part of a non-white racial or ethnic group. Also due to the lack of adequate sample sizes, we were unfortunately unable to explore the transportation needs of children identifying as members of the LGBTQ+ community or with different ability statuses. There may be important differences in the nature of the transportation needs or need resolutions among these populations served by public behavioral health systems; better understanding the transportation needs and impact of transportation barriers within these populations is another crucial area for future work.

Second, we used a single item assessment of transportation needs due to the nature of the CANS instrument. The CANS is designed for breadth of assessment, and, therefore, it will not fully capture the depth of any particular area of assessment. These data do not capture the nature of the transportation need (e.g., was the transportation need due to not having a personal vehicle or due to limited access to public transportation?) or the nature of the need resolution (e.g., was the need resolved due to direct therapeutic intervention or did the caregiver resolve this need on their own?). In the future, researchers may consider investigating the magnitude of effects of transportation need resolution based on the nature of the need itself and how it is resolved.

Finally, this study was observational in nature. As such, we were not able to delineate a mechanism of change from these data to explain why the relationship between transportation need resolution and the resolution of other needs/strength occurred in this population. It is possible that transportation needs increased clients’ access to healthcare specifically, increased engagement in services and activity more broadly, allowed clients to better build relationships and community, or any number of other possibilities. It is furthermore possible that the observed resolution of transportation needs was simply an indicator of increased financial stability for the family. While we did control for caregiver financial needs in this analysis, we cannot definitively rule out the possibility that transportation needs are a proxy for overall financial resources. As this study was observational, it will be important for future studies to replicate these findings and establish a causal nature among the relationships between caregiver transportation needs, need resolution, and the downstream impact on child-level outcomes.

More work is required to better understand the nature of transportation need resolution, particularly to develop the literature around how transportation needs impact children’s health and healthcare. What is the typical length of time it takes to resolve transportation needs such that they are no longer a barrier to improvement in other areas? How does the rate and magnitude of need resolution vary based on client demographic variables, geographic location and population density of the client’s environment, or other needs of the caregiver? The results of this study demonstrate that the resolution of transportation needs has a large downstream impact on children’s psychosocial functioning across multiple domains, but we were not able to investigate whether transportation need resolution was particularly impactful on certain CANS items or areas of functioning. Investigations of this kind may lead to additional methodological advances for the study of complex assessment and systems, allowing investigators to weight items within larger predictive models to gain a more nuanced understanding of change and item impact over time.

Despite the limitations of this project, we see this study as an important contribution to the literature on the impact of caregiver transportation needs on child-level outcomes, particularly since the outcomes used in this study represent a broad range of children’s psychosocial needs and strengths. We hope that future work will expand upon the findings of this study to investigate the relationships between transportation needs and children’s global functioning in more diverse populations, such as for children identifying as part of a non-white racial or ethnic groups, as members of the LGBTQ+ community, or as having different ability statuses. We also hope that future investigators are able to replicate the findings of this study to establish causal relationships between caregiver transportation needs and child-level outcomes to ultimately aid in administrative and clinical decision support.

### 4.2 Conclusion

The results of this study are the first to explore the impact of caregiver transportation needs on children’s broad psychosocial functioning and change over the course of care within a large, public behavioral health system. We demonstrated not only that caregiver transportation needs meaningfully differentiate children at the beginning of treatment, but also that the resolution of caregiver transportation needs has a far-reaching and positive downstream impact for children. Many research questions are still outstanding, particularly as we continue to redefine how healthcare access is understood for children and families served in complex systems. In light of the COVID-19 pandemic, the nature of healthcare access and the meaning of “transportation” has changed quickly, dramatically, and possibly permanently. Many administrators, clinicians, and researchers have recently focused on issues of internet access as important drivers of healthcare services, particularly as delivery of services via telehealth has become more common in 2020. We would suggest that for many children and families, unmet transportation needs will still have far-reaching impacts on a child’s psychosocial functioning, even if access to healthcare is in some ways improved by telehealth. It will be important to replicate the findings of this study within a population of children served during the COVID-19 pandemic in order to understand how transportation needs have impacted care, and which children and families were still profoundly impacted by transportation needs despite the changing landscape of healthcare delivery.

Based on the results of this study, it appears that the resolution of transportation needs is a critical area of clinical intervention for children served in public behavioral and mental healthcare systems. It is our hope that these results will be generative of future work around understanding how best to resolve transportation needs in this population. In addition, we hope that the importance of resolving transportation needs is highlighted for clinicians and administrators working in complex systems that serve children and families. On an individual level, this work may impact treatment planning decisions: prioritizing the resolution of transportation needs may make it easier to resolve psychosocial needs in other areas. On a systems level, administrators of complex systems that serve children and families may consider additional resource allocation dedicated to understanding and resolving transportation needs of their clients as a way to enhance treatment success throughout the system.

## Data Availability

The data that support the findings of this study are available from the Idaho Department of Behavioral Health ICANS database, but restrictions apply to the availability of these data, which were used under license for the current study, and so are not publicly available.

